# A territory-wide study of arrhythmogenic right ventricular cardiomyopathy patients from Hong Kong

**DOI:** 10.1101/2021.11.17.21266304

**Authors:** Ishan Lakhani, Jiandong Zhou, Sharen Lee, Ka Hou Christien Li, Keith Sai Kit Leung, Guoliang Li, Tong Liu, Wing Tak Wong, Ian Chi Kei Wong, Ngai Shing Mok, Chloe Mak, Qingpeng Zhang, Gary Tse

## Abstract

**Background:** Arrhythmogenic right ventricular cardiomyopathy/dysplasia (ARVC/D) is a hereditary disease characterized by fibrofatty infiltration of the right ventricular myocardium that predisposes affected patients to malignant ventricular arrhythmias, dual-chamber cardiac failure and sudden cardiac death (SCD). The present study aims to investigate the risk of detrimental cardiovascular events in an Asian population of ARVC/D patients, including the incidence of malignant ventricular arrhythmias, new-onset heart failure with reduced ejection fraction (HFrEF), as well as long-term mortality.

**Methods and Results:** This was a territory-wide retrospective cohort study of patients diagnosed with ARVC/D between 1997 and 2019 in Hong Kong. This study consisted of 109 ARVC/D patients (median age: 61 [46-71] years; 58% male). Of these, 51 and 24 patients developed incident VT/VF and new-onset HFrEF, respectively. Five patients underwent cardiac transplantation, and 14 died during follow-up. Multivariable Cox regression identified prolonged QRS duration as a predictor of VT/VF (p < 0.05). Female gender, prolonged QTc duration, the presence of epsilon waves and T-wave inversion (TWI) in any lead except aVR/V1 predicted new-onset HFrEF (P < 0.05. The presence of epsilon waves, in addition to the parameters of prolonged QRS duration and worsening ejection fraction predicted all-cause mortality (p<0.05). Clinical scores were developed to predict incident VT/VF, new-onset HFrEF and all-cause mortality, and all were significantly improved by machine learning techniques.

**Conclusion:** Clinical and electrocardiographic parameters are important for assessing prognosis in ARVC/D patients and should in turn be used in tandem to aid risk stratification in the hospital setting.

## Introduction

Arrhythmogenic right ventricular cardiomyopathy/dysplasia (ARVC/D) is a rare hereditary condition presenting at an incidence of 1 in 2500 to 1 in 5000 in the general population, with notable geographical variations in disease prevalence (1). ARVC/D is characterized by genetic mutations in desmosomal genes (2,3) and accompanying aberrations in cardiomyocyte cell-cell adhesion, leading to early cardiac regional anatomical abnormalities, typically confined to the right ventricular inflow tract, outflow tract, and apex, which together constitute the “triangle of dysplasia” (4). Disease progression is in turn dominated by diffuse thinning of the right ventricular wall with cardiomyocyte loss and corresponding fibrofatty replacement of the myocardium (5). These pathological alterations not only disturb the native electrical conduction system, thereby predisposing affected patients to malignant ventricular arrhythmias and sudden cardiac death (SCD) (6,7), but also potentially induce left ventricular dysfunction and subsequent dual chamber cardiac failure (8).

The definitive diagnosis of ARVC/D is challenging owing to the absence of a single set of parameters sufficiently specific to the disease (1). As such, the current diagnostic criterion seeks to amalgamate a series of clinical, pathological and genetic features most commonly observed in affected patients, amongst which electrocardiographic and echocardiographic parameters are the most prominent (9). The evident heterogeneity in the phenotypic presentation and complications associated with ARVC/D poses difficulties to optimal management (10). Current therapies are primarily geared towards the prevention of lethal ventricular arrhythmias, and implantable cardioverter-defibrillator (ICD) placement has hitherto proven to be the only effective strategy in reducing long-term mortality (1). Such dilemmas in the management of these patients are further compounded by the apparent underreporting of prognostic markers to assist risk stratification in the clinical setting.

The present study aims to investigate the risk of detrimental cardiovascular events in an Asian population of ARVC/D patients, including the incidence of malignant ventricular arrhythmias, new-onset heart failure with reduced ejection fraction (HFrEF), as well as long-term mortality. Moreover, the prognostic importance of several clinical parameters will be examined in an attempt to identify possible markers with predictive value that could improve overall assessment and therapeutic guidance.

## Methods

### Diagnosis of ARVC/D

In 1994, an International Task Force (ITF) proposed an initial criterion for ARVC/D recognition, based on six major categories: i) global and/or regional dysfunction and structural alterations of the right ventricle, ii) tissue characterization of the right ventricular wall, iii) repolarization abnormalities, iv) depolarization abnormalities, v) cardiac arrhythmias, and vi) family history. Each category comprised of one or more major and/or minor requirements, from which several different permutations of major and minor variable combinations were considered diagnostic of ARVC/D. With time, the discovery of new associated histological, electrocardiographic and echocardiographic parameters with greater sensitivity for the detection of early-stage disease led to the proposition of the revised ITF criteria in 2010. The modified criteria elaborated upon the initial guidelines in greater detail, the specifics of which can be found elsewhere (11).

### Study population and their baseline characteristics

This study was approved by The Joint Chinese University of Hong Kong – New Territories East Cluster Clinical Research Ethics Committee. The current study included ARVC/D patients who presented to public hospitals managed by the Hospital Authority of Hong Kong between January 1999 to December 2019. Patient data was obtained using the Clinical Management System (CMS), an electronic health database that is connected to the territory-wide Clinical Data Analysis and Reporting System (CDARS). Both systems are integrative centralized platforms that permit the extraction of clinical data for analysis and reporting. The collaborative use of CMS and CDARS systems allowed for the retrieval of comprehensive medical records, including disease diagnoses, clinical comorbidities, electrocardiographic indices, echocardiographic parameters and operative procedures. Our teams have used these systems for studying other ion channelopathies in the territory (12,13). In the present study, ARVC/D subjects were recruited by International Statistical Classification of Diseases (ICD) coding and with subsequent diagnostic confirmation by cardiologist review. Collected patient data included: 1) age, 2) gender, 3) age at ARVC/D diagnosis, 4) family history of ARVC/D and VF/SCD, 5) presentation of palpitations, 6) presentation of syncope and the number of episodes, 7) presentation of premature ventricular contractions (PVCs) and PVC burden, 8) pre-existing ventricular tachycardia / ventricular fibrillation (VT/VF) prior to ARVC/D diagnosis and the number of episodes,9) incident non-sustained ventricular tachycardia (NSVT) and the number of episodes, 10) performance of electrophysiological study (EPS) and presentation of EPS-induced VT/VF, 11) performance of 24-hour ECG Holter, 12) performance of theexercise stress test, 13) ICD implantation, and 14) operative heart transplantation.

Further data collection involved using these electronic databases to obtain echocardiographic reports closest to the date of ARVC/D diagnosis in order to determine left ventricular ejection fractions (LVEF) and confirm the presence of right ventricular morphological pathologies consistent with ARVC/D diagnosis, including right ventricular dyskinesia, dilatation, aneurysms, fibrofatty replacement and systolic dysfunction. Likewise, automated electrocardiogram (ECG) recordings taken closest to the date of ARVC/D diagnosis were also extracted for the following indices: 1) ventricular rate, 2) P-wave duration, 3) PR-interval, 4) QRS duration, 5) QT and QTc interval, 6) T-wave inversion, 7) R-wave amplitude in V5, 8) S-wave amplitude in V1, 9) manifestation of epsilon waves, 10) P-wave axis: representing the net vectorial direction of atrial depolarization, 11) QRS axis and T-wave axis: representing the net vectorial depolarization and repolarization, respectively. Moreover, the primary long-term outcome assessed was incident VT/VF post-ARVC/D diagnosis. Secondary outcomes derived included: 1) new-onset HFrEF) defined as LVEF ≤40%, and 2) all-cause mortality.

### Statistical analysis

Descriptive statistics were presented as median [interquartile range] or as count (percentage) as appropriate. The study population was stratified according to the presence or absence of incident VT/VF. The Mann-Whitney U test was used to compare continuous variables. Chi-squared test with Yates’ correction was used for 2×2 contingency data, and Pearson’s Chi-squared test was used for contingency data for variables with more than two categories. The relationship between electrocardiographic and clinical parameters with outcomes were assessed using univariable Cox proportional-hazards model. Variables with P<0.05 were incorporated into a multivariable model, as well as a scoring system. Briefly pertaining to the scoring system, a point assigned to a variable was equivalent to the halved value of the hazard ratio, rounded up to the nearest integer. Statistical analysis were performed using Stata (Version 13.0). A two-sided P-value < 0.05 was considered statistically significant.

### Development of a machine learning survival learning model

A non-parametric machine learning survival analysis model was developed to predict incident VT/VF, new-onset HFrEF and all-cause mortality in ARVC/D patients. The underlying motivation for the implementation of machine learning survival analysis models stemmed from the ability of these algorithms to better capture nonlinear and interactive patterns within survival data compared to traditionally used Cox regression models, which assume the existence of a hazard function between survival data and censored outcomes. A major problem pertaining to the use of Cox regression models is the assumption of a linear relationship between covariables and the time of event occurrence. Many modifications have been proposed aiming to circumvent this limitation, namely by generalizing the Cox regression model to take into account the corresponding non-linear and interactive relations between covariables and the time of event. Survival trees (14) and random survival forests (RSF) (15) models were developed on the premonition that tree-based models, after being combined with baseline models (e.g. decision trees), can generate the best survival predictions. Recently, a weighted random survival forests (wRSF) (16) model was proposed as an efficient modification of RSF models by replacing the standard procedure of averaging used for the estimation of RSF hazard function with a weighted averaging strategy, wherein the weights are assigned to every tree and can be viewed as training parameters computed by maximizing Harrell’s concordance index (C-index).

The present study introduced a wRSF model for prediction of incident VT/VF, HFrEF and all-cause mortality after first presentation of ARVC/D. The most important variables for outcome prediction were derived with a variable importance ranking approach of the wRSF model. The ranked results were subsequently used to construct a machine learning-based electronic frailty index with prognostic value in assessing the incidence of the three aforementioned outcomes. The survival prediction performance of wRSF, RSF and multivariable Cox models in discriminating incident VT/VF, HFrEF and all-cause mortality were compared using several evaluation measures, including precision, recall, area under the receiver operating characteristics curve (AUC), and Harrell’s C-index. The comparative experiments were conducted based on the input of significant univariable predictors identified by the initial univariable Cox proportional-hazards model. R packages, including *randomForestSRC*(Version 2.1.5), *randomForestSRC* (Version 2.9.3), *survival* (Version 2.42-3) and *ggplot2* (Version 3.3.2), were used to generate the survival prediction results.

## Results

In this ARVC/D cohort (n = 109), the median age was 61 [46-71] years and 63 (58%) were male. The baseline characteristics are presented in **Table 1**, with patients stratified according to the development of incident VT/VF. The median ages of the VT/VF (n = 49) and non-VT/VF (n = 60) groups were 65 [45-71] years and 59 [46.5-71.5] years, respectively with similar ages at diagnosis of ARVC/D. Patients who developed incident VT/VF tended to present more often with right ventricular dilatation and systolic dysfunction. This group also demonstrated a significantly longer QRS duration, which took a median value of 108.0 [96.0-129.0]ms, as well as a significantly greater proportion of subjects developing epsilon waves (n = 13; 27%). Anti-arrhythmic therapy was prescribed in the form of amiodarone (n = 48), sotalol (n = 34) and mexiletine (n = 4), with majority of patients taking these medications after the first episode of VT/VF (amiodarone: n = 28; sotalol: n = 21; mexiletine: n = 4). A total of 59 patients received implantable cardioverter-defibrillator (ICD) placement either for VT/VF prophylaxis following ARVC/D diagnosis (n = 33) or to prevent VT/VF recurrence after the first episode (n = 26).

**Table 1.**
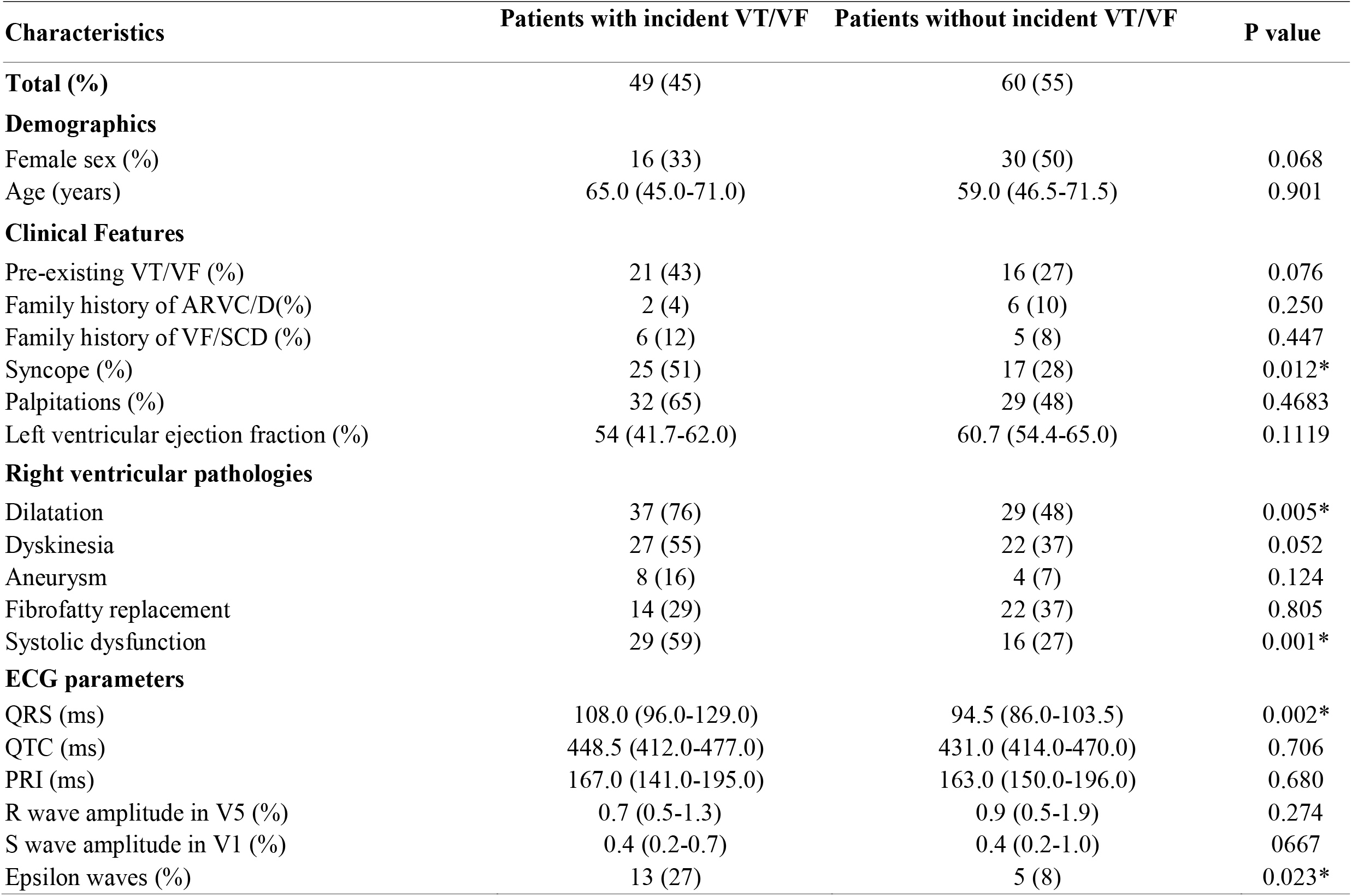
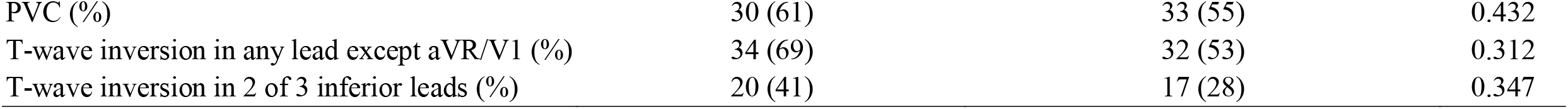
Baseline characteristics for patients with and without incident VT/VF.

### Predictors of adverse outcomes

In the following ARVC/D cohort, 49 patients and 24 patients developed incident VT/VF and new-onset HFrEF, respectively. A total of 5 patients underwent cardiac transplantation, and 14 patients passed away during follow-up, 11 of which suffered from ARVC/D-related complications, namely SCD and HFrEF, whereas the remaining 3 suffered non-cardiovascular related deaths. The results of univariable Cox proportional-hazards regression analysis for predicting incident VT/VF, new-onset HFrEF and all-cause mortality are reported in **Supplementary Tables 1, 2 and 3**, respectively, with the corresponding Kaplan-Meier survival curves shown in **Figure 1**.

**Figure 1:**
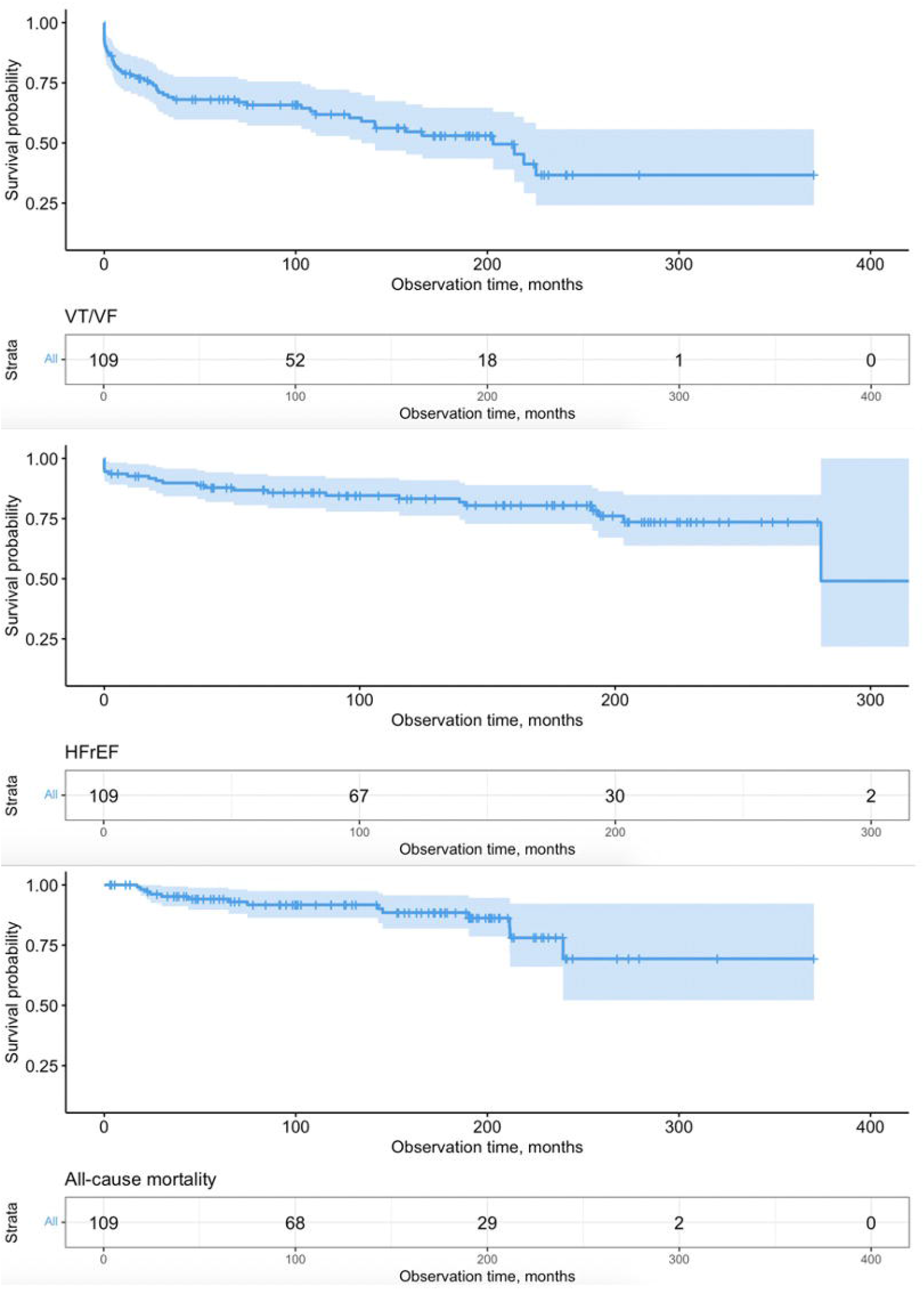
Kaplan-Meier survival curves for incident VT/VF, new-onset HFrEF and all-cause mortality.

Incident VT/VF **(Supplementary Table 1)** was associated with prolonged QRS duration, presence of epsilon waves, and syncope, with only the foremost retaining significance after multivariable adjustment (p<0.05). Regarding secondary outcomes, univariable predictors of new-onset HFrEF **(Supplementary Table 2)** included female gender, prolonged QTc duration, presence of epsilon waves and TWI in any lead except aVR/V1, all of which retained significance in the multivariable model (p<0.05). Likewise, all-cause mortality **(Supplementary Table 3)** was similarly associated with all univariable predictors of new-onset HFrEF, in addition to prolonged QRS duration (p<0.05). Resultant significant parameters in multivariable Cox regression included female gender, prolonged QRS duration, prolonged QTc duration, and presence of epsilon waves (p<0.05).

### Scoring system for new-onset VT/VF in ARVC/D

Significant clinical and electrocardiographic parameters in univariable Cox regression (P < 0.05) were used to design a scoring system to predict new-onset VT/VF in ARVC/D. Receiver operator characteristics (ROC) curves were used to determine optimal cut-offs for significant continuous variables. The optimal cutoff value for QRS duration was 98.5ms (AUC: 0.69; sensitivity = 72%; specificity = 67%). After categorization, this parameter retained significance in univariable prediction of incident VT/VF in ARVC/D, and was therefore eligible for inclusion. QRS duration > 98.5ms, along with presence of syncope and epsilon waves were subsequently used to form the final scoring system (**Supplementary Table 4a)**. Subjects who developed VT/VF presented with a median score that was 0.5 points higher than those who remained free of VT/VF. Cox proportional-hazards analysis revealed that patients with a per unit increase in the score had a 74% higher risk of incident VT/VF (HR: 1.74; 95% CI: 1.30-2.33; P < 0.001) **(Supplementary Table 4b)**. Categorization of the VT/VF score using the maximal rank statistics approach **(Figure 2)** revealed an optimal cut-off of 1 point. Subsequent Cox proportional-hazards analysis demonstrated that patients a score ≥1 point had an approximate 2-fold increase in risk of new-onset VT/VF **(Supplementary Table 4c)**.

**Figure 2:**
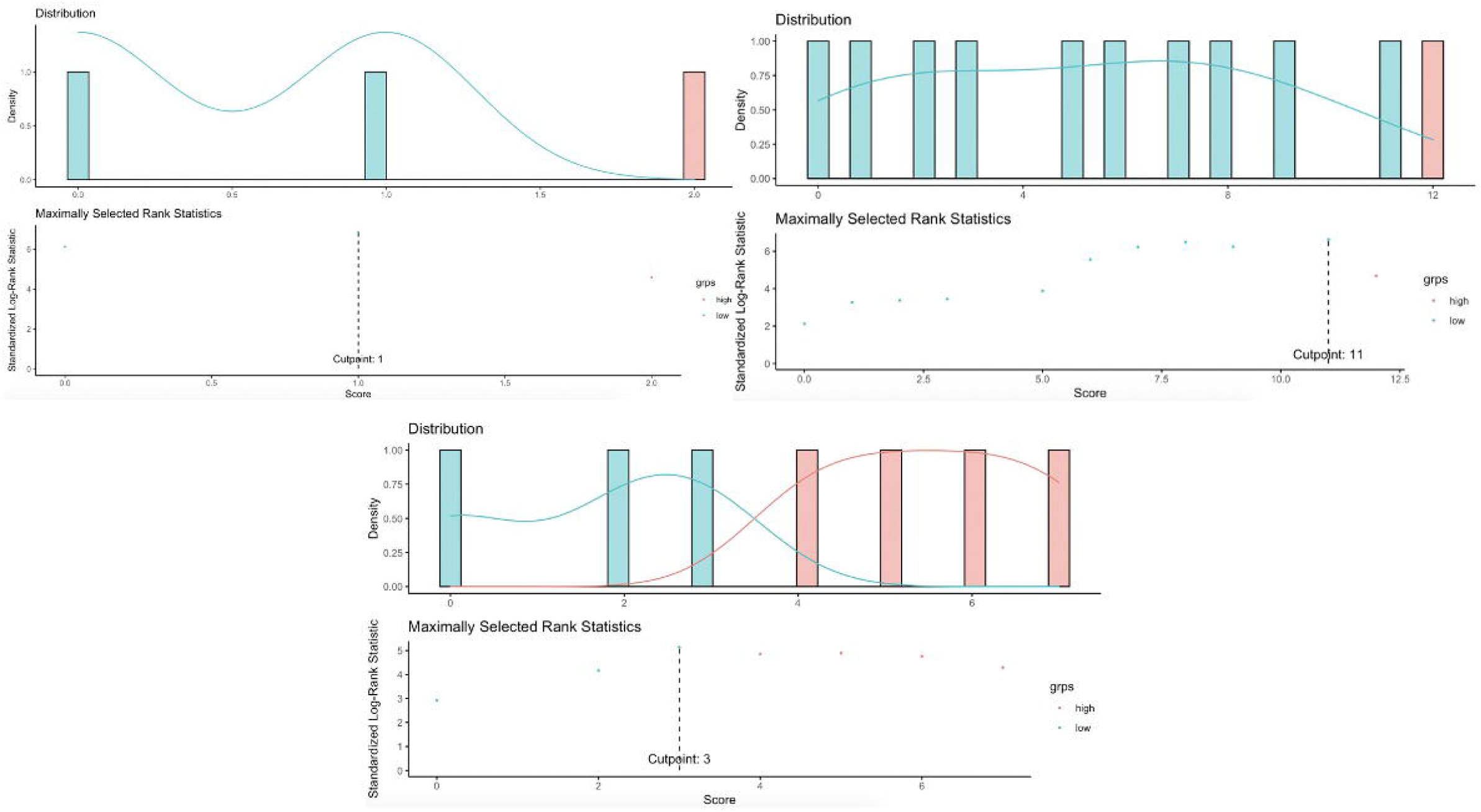
Optimal cut-off for the scoring system for incident VT/VF, new-onset HFrEF and all-cause mortality with the maximal rank statistics approach.

### Scoring system for new-onset HFrEF in ARVC/D

A similar process was conducted for the creation of a score for HFrEF. The optimal cutoff value for QTc duration was 437.5ms (AUC: 0.76; sensitivity = 88%; specificity = 65%), which remained significant in univariable prediction of VT/VF in ARVC/D. Overall, four binary parameters were ultimately included in the final scoring system, including female gender, presence of epsilon waves, presence of TWI in any lead except aVR/V1, and QTc > 437.5ms **(Supplementary Table 5a)**. Patients with HFrEF presented with a median score that was 6 points higher compared to those without HFrEF **(Supplementary Table 5b)**. Cox proportional-hazards analysis revealed that patients with a higher score, when used as a continuous variable, had a 48% higher risk of new-onset HFrEF (HR: 1.48; 95% CI: 1.27-1.74; P < 0.001) **(Supplementary Table 5c)**. Likewise, categorization of the HFrEF score using the maximal rank statistics approach **(Figure 2)** revealed an optimal cut-off of 11 points, using which it was shown that those with a score ≥11 points had a more than 15-fold increase in risk of new-onset HFrEF **(Supplementary Table 5c)**.

### Scoring system for all-cause mortality in ARVC/D

As it pertains to all-cause mortality, the optimal cutoff values for i) QRS duration was 122.5ms (AUC = 0.71; sensitivity = 57%; specificity = 85%) and ii) QTc duration was 448.5ms (AUC = 0.73; sensitivity = 79%; specificity = 63%), both of which retained significance in univariable prediction of all-cause mortality in ARVC/D. Per unit decrease in LVEF was also a predictor of all-cause mortality, but failed to retain significance following categorization and was therefore not included in the scoring system. All in all, a total of four binary parameters were subsequently included in the final scoring system, including gender, presence of epsilon waves, QRS > 122.5ms, and QTc > 448.5ms **(Supplementary Table 6a)**. Patients who suffered death presented with a median score that was 4.5 points higher compared to those who survived **(Supplementary Table 6b)**. Cox proportional-hazards analysis revealed that patients with a per unit increase in the score had a 65% higher risk of all-cause mortality (HR: 1.65; 95% CI: 1.33-2.06; P < 0.001) **(Supplementary Table 6c)**. With an optimal cut-off of 3 points determined by the maximal rank statistics approach **(Figure 2)**, those with a score ≥3 points had an almost 23-fold increase in risk of mortality **(Supplementary Table 6c)**.

### Machine learning to predict incident VT/VF, new-onset HFrEF and all-cause mortality

wRSF models were used to predict primary and secondary outcomes with the identified significant parameters in univariable analysis as input variables (p value<0.05). The optimal tree number used to build each wRSF model was selected by the error rate minimization with iteration approach. The tree number selected to predict VT/VF, new-onset HFrEF and all-cause mortality was 250, 200, and 400 respectively (**Figure 3**). The derived importance value ranking of the variables are shown in **Table 2**. QRS duration > 98.5ms was the most predictive variable for incident VT/VF, followed by presence of syncope and presence of epsilon waves. In contrast, prolonged QTc duration demonstrated the strongest predictivity for HFrEF, followed by TWI in any lead except aVR/V1, presence of epsilon waves, and female gender. As it pertains to all-cause mortality, QRS > 122.5ms was the most important predictor, followed by presence of epsilon waves, QTc > 448.5ms, and female gender. As shown in **Table 3**, the ability of the wRSF model to predict the primary outcomes was compared with an RSF model and multivariable Cox model, based on evaluation metrics of precision, recall, AUC and C-index. Findings indicate that the wRSF models performed best in the prediction of all three outcomes based on the significant univariable predictors.

**Figure 3:**
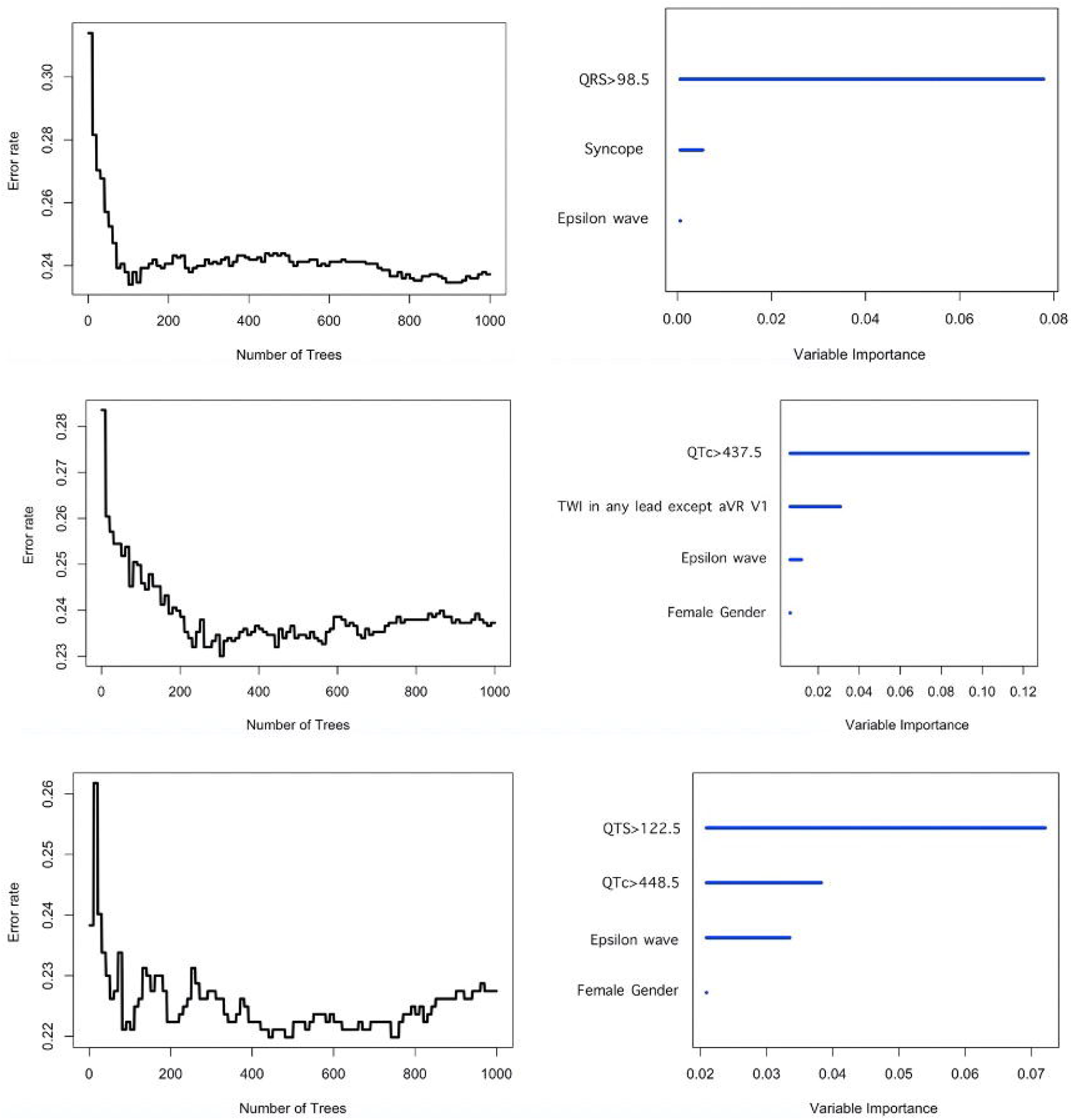
Optimal tree number of wRSF model and variable importance ranking to predict incident VT/VF, new-onset HFrEF and all-cause mortality (bottom).

**Table 2.**
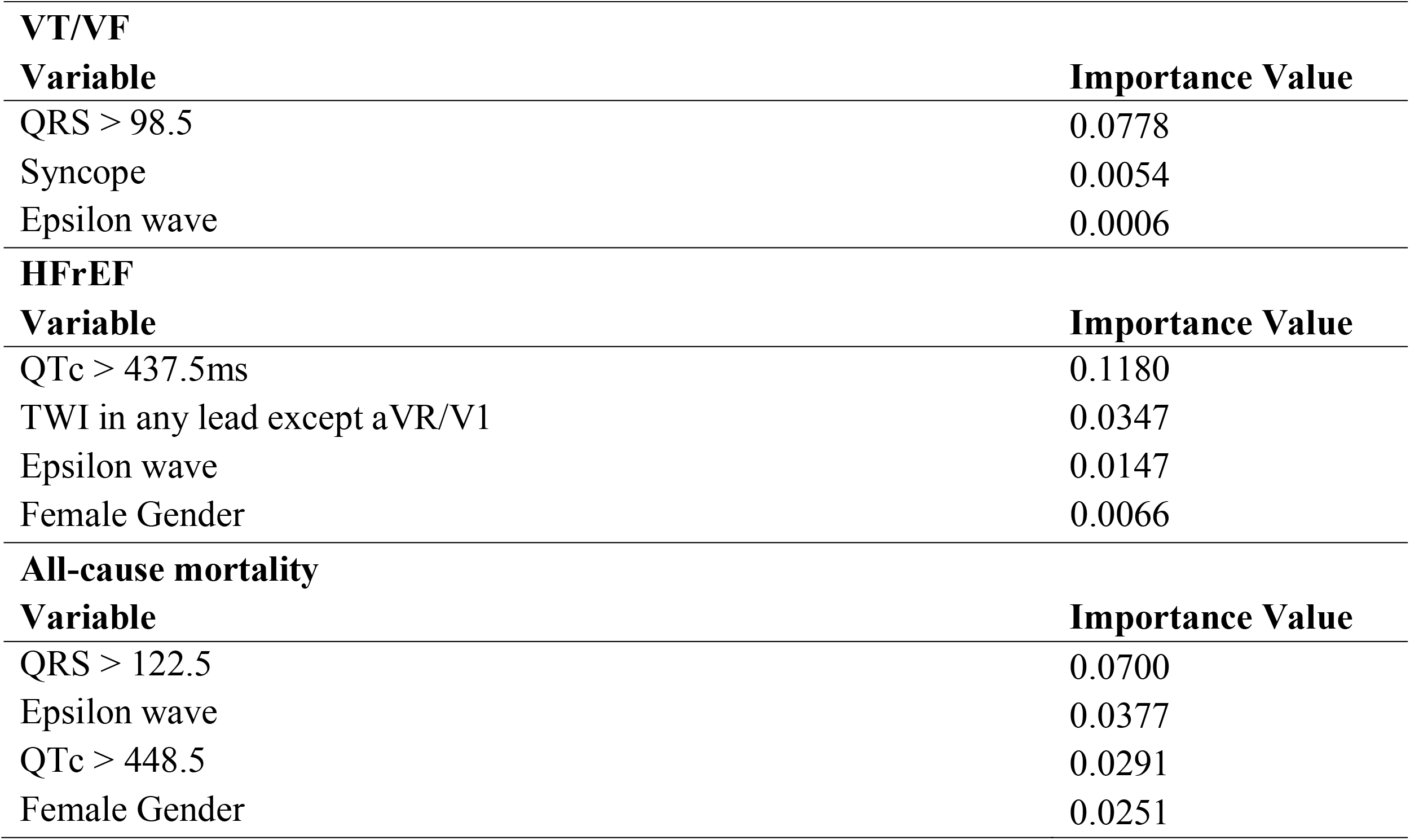
Variable importance ranking generated by wRSF models for primary and secondary outcomes.

| **VT/VF** |  |
| --- | --- |
| **Variable** | **Importance Value** |
| QRS > 98.5 | 0.0778 |
| Syncope | 0.0054 |
| Epsilon wave | 0.0006 |
| **HFrEF** |  |
| **Variable** | **Importance Value** |
| QTc > 437.5ms | 0.1180 |
| TWI in any lead except aVR/V1 | 0.0347 |
| Epsilon wave | 0.0147 |
| Female Gender | 0.0066 |
| **All-cause mortality** |  |
| **Variable** | **Importance Value** |
| QRS > 122.5 | 0.0700 |
| Epsilon wave | 0.0377 |
| QTc > 448.5 | 0.0291 |
| Female Gender | 0.0251 |

**Table 3.** Performance comparison between wRSF, RSF, and multivariable Cox model.

| **Incident VT/VF** |  |  |  |  |
| --- | --- | --- | --- | --- |
| **Model** | **Precision** | **Recall** | **AUC** | **C-index** |
| wRSF model | <u>0.8352</u> | <u>0.8478</u> | <u>0.8341</u> | <u>0.8202</u> |
| RSF model | 0.8283 | 0.8372 | 0.8161 | 0.8116 |
| Multivariable Cox model | 0.7493 | 0.7793 | 0.7524 | 0.7835 |
| **New-onset HFrEF** |  |  |  |  |
| **Model** | **Precision** | **Recall** | **AUC** | **C-index** |
| wRSF model | <u>0.8290</u> | <u>0.8305</u> | <u>0.8363</u> | <u>0.8182</u> |
| RSF model | 0.8053 | 0.8172 | 0.8184 | 0.8021 |
| Multivariable Cox model | 0.7493 | 0.7641 | 0.7620 | 0.7770 |
| **All-cause mortality** |  |  |  |  |
| **Model** | **Precision** | **Recall** | **AUC** | **C-index** |
| wRSF model | <u>0.7322</u> | <u>0.7395</u> | <u>0.7549</u> | <u>0.7481</u> |
| RSF model | 0.7071 | 0.6922 | 0.6984 | 0.7012 |
| Multivariable Cox model | 0.6734 | 0.6808 | 0.6853 | 0.6744 |

## Discussion

The present study is novel as it is among the first territory-wide investigations of ARVC/D patients in Hong Kong, allowing for the development of the first clinical risk scores for predicting incident VT/VF, new-onset heart failure as well as mortality in ARVC/D. In addition, the application of machine learning algorithms to assess ARVC/D prognosis has also not previously been investigated, and in turn demonstrated enhanced risk prediction for outcomes compared to other analytical models.

The substrate for arrhythmogenesis in ARVC/D is a combination of conduction and repolarization abnormalities associated with structural alterations in the right ventricle (17). However, recent work has demonstrated that non-classical forms of arrhythmogenic cardiomyopathy, namely left dominant or biventricular forms, are more prone to ventricular arrhythmias than classical ARVC/D (18). Moreover, the atria, in addition to the ventricles, are also abnormal in ARVC/D (19) with complications such as sinoatrial arrests and atrial fibrillation (20,21). Different ECG indices have been identified as risk markers of ventricular arrhythmias (22), amongst which the epsilon wave is the classical pathognomonic feature of ARVC/D (23,24). Repolarization criteria, including TWI in inferior leads, a precordial QRS amplitude ratio of ≤ 0.48, and QRS fragmentation also constitute valuable variables for predicting adverse outcomes in this disease (25). Repolarization abnormalities, such as TWI, are important, and electroanatomic mapping areas have shown to be proportional to extent of TWI on 12-lead ECG (26) (27). Strain imaging by speckle-tracking echocardiography has been used to risk-stratify patients in heart failure (28) and recent work has shown that incorporation of mechanical dispersion can further improve risk prediction (29), as heart failure is typically under-recognized in ARVC/D (30).

Ventricular arrhythmias are a common occurrence amongst patients with so-called inherited cardiac arrhythmias, including ARVC/D, Brugada syndrome, long QT-syndrome and short QT-syndrome, amongst which ARVC/D cohorts have shown to present with the highest rates of frequent ventricular premature complexes, (non)sustained ventricular tachycardias, and malignant ventricular tachyarrhythmias (31). Several large-scale studies have reported on the clinical characteristics and predictors of ventricular arrhythmias in ARVC/D patients. In 131 definite ARVC/D patients, spontaneous sustained ventricular arrhythmias, cardiac syncope, male gender, proband, and inducibility in electrophysiology study were all significant predictors of incident sustained VT/VF and SCD (32). The same group further studied the phenotype of ARVC/D patients with late presentation, demonstrating that this subpopulation does not confer a benign prognosis and has a high arrhythmic risk (33). Another study of 135 patients identified prolonged QRS duration on signal-averaged ECG, non-sustained VT on 24 h-ECG, and the absence of negative T waves in lead aVR on a 12-lead surface ECG as significant predictors of recurrent sustained ventricular arrhythmias and hospitalization due to ventricular arrhythmias in ARVC/D (34). Moreover, an investigation of 137 patients from France found that low LVEF, positive electrophysiological studies and physical activity >6 h/week were shown to be independently associated with the development of ventricular arrhythmias (35). The findings of the aforementioned studies clearly demonstrate the high incidence of ventricular arrhythmias within ARVC/D cohorts, thereby necessitating the use of prophylactic antiarrhythmics to reduce their occurrence (36), along with radiofrequency catheter ablation for patients who end up developing these rhythm abnormalities(37).

Recently, a systematic review and meta-analysis summarized the current literature, identifying consistently predictive risk factors in patients with definite ARVC across different studies (38). These were male sex, syncope, TWI in lead V3, right ventricular dysfunction, and prior (non)sustained VT/VF. Our present work extends these findings by demonstrating that longer QRS duration, the presence of epsilon waves and TWI in 2/3 inferior leads were significantly associated with incident VT/VF, albeit only QRS remained a significant predictor after multivariable adjustment. Several parameters retained significance in multivariable prediction of new-onset HFrEF, including longer QTc duration, presence of epsilon waves, TWI in any lead except aVR/V1 and female gender. Likewise, longer QRS duration, presence of epsilon waves, LVEF and age at diagnosis of ARVC/D were all significantly associated with all-cause mortality in multivariable analysis. It was then possible to further enhance risk prediction through the application of wRSF model analysis, which we have recently used to better risk prediction in acquired long QT syndrome (39) as well as Brugada syndrome (40). The wRSF model was able to improve the risk stratification for incident VT/VF, new-onset HFrEF and all-cause mortality in this ARVC/D cohort.

Furthermore, the clinical heterogeneity typically observed amongst populations with ARVC/D makes the use of scoring algorithms a potentially useful method to amalgamate the different patient parameters for the purposes of risk stratification. Such an approach has been adopted previously in a large cohort of 528 ARVC/D patients to predict the long-term risk of ventricular arrhythmias. The model constructed, which included age, male gender, cardiac syncope in the prior 6 months, prior non-sustained VT, number of PVCs in 24 h, number of leads with TWI and right ventricular ejection fraction, demonstrated an improved ability to estimate risk of ventricular arrhythmias and guide decision-making in ICD implantation for such patients (41). A meta-analysis identified the following 11 variables as the most important factor for predicting arrhythmic events: (1) male gender, (2) presyncope, (3) left ventricular dysfunction, (4) TWI in inferior leads, (5) proband status, (6) late potentials, (7) syncope, (8) inducibility at electrophysiological study, (9) right ventricular dysfunction, (10) epsilon waves, and (11) premature ventricular contractions greater than 1000/24 h (42). To our knowledge, such scoring algorithms have not been used to investigate outcomes beyond VT/VF in ARVC/D cohorts. As such, the present study also developed two multi-parametric scores for predicting new-onset HFrEF and all-cause mortality, respectively, both of which demonstrated efficacy in assessing ARVC/D patient prognosis.

## Limitations

This investigation has limitations that should be noted. Firstly, data is primarily based on patients of Chinese ethnicity and therefore lacks the subject variability needed for a comprehensive evaluation of ARVC/D, which itself presents with a heterogeneous phenotype. Secondly, several subjects were prescribed amiodarone and/or sotalol as treatment, both of which have been previously shown to lengthen QTc interval. This could have potentially influenced the reported relationship between QTc duration and new-onset HFrEF as well as all-cause mortality. Finally, the adverse ECG findings, such as prolongation of QRS duration or QTc duration and the presence of epsilon waves, are likely linked to a greater underlying disease severity that in turn leads to malignant ventricular arrhythmias. As such, these parameters possibly only serve as markers, as opposed to outright predictors, of VT/VF, albeit further study is required to confirm this.

## Conclusions

The phenotypic variability and adverse prognosis potentially associated with ARVC/D necessitates a multimodality approach for risk stratification that includes both clinical and electrocardiographic parameters. Combinatorial methods involving machine learning algorithms are able to account for underlying inter-variable interactions, thereby improving overall event and survival prediction. As a result, with further study into their use, machine learning techniques could possibly provide an alternative, more effective avenue to assess patient prognosis in such heterogeneous disease conditions.

## Data Availability

All data produced in the present study are available upon reasonable request to the authors

## Figure Legends

**Supplementary Table 1.** Cox proportional-hazards model to predict incident VT/VF.

|  | Incident VT/VF |  |
| --- | --- | --- |
|  | Univariable Hazards Ratio<br>(P value; 95% CI) | Multivariable Hazards Ratio<br>(P value; 95% CI) |
| Female gender | 0.66 (0.36-1.19; P = 0.170) |  |
| Family history of ARVC/D | 0.49 (0.12-2.03; P= 0.328) |  |
| Family history of VF/SCD | 1.16 (0.79-2.49; P = 0.737) |  |
| Pre-existing VT/VF | 1.44 (0.81-2.55; P = 0.210) |  |
| Syncope | **1.83 (1.04-3.23; P = 0.036)** | 1.27 (0.68-2.37; P = 0.76) |
| Palpitations | 1.77 (0.97-3.22; P = 0.065) |  |
| LVEF | 0.98 (0.96-1.00; P = 0.117) |  |
| QRS duration | **1.02 (1.008-1.03; P < 0.001)** | **1.01 (1.0002-1.02; P = 0.013)** |
| QTc duration | 1.00 (0.99-1.01; P=0.330) |  |
| PR interval | 1.00 (0.99-1.003; P = 0.400) |  |
| R wave amplitude in V5 | 0.63 (0.34-1.15; P = 0.169) |  |
| S wave amplitude in V1 | 0.79 (0.32-1.99; P = 0.719) |  |
| Epsilon waves | **2.34 (1.22-4.48; P = 0.011)** | 1.46 (0.83-3.64; P = 0.144) |
| PVC | 1.18 (0.65-2.12; P = 0.583) |  |
| TWI in any lead except aVR/V1 | 1.53 (0.81-2.92; P = 0.188) |  |
| TWI in 2/3 inferior leads | 1.77 (0.984- 4.48; P = 0.057) |  |

**Supplementary Table 2.** Cox proportional-hazards model to predict new-onset HFrEF.

|  | New-onset HFrEF |  |
| --- | --- | --- |
|  | Univariable Hazards Ratio<br>(P value; 95% CI) | Multivariable Hazards Ratio<br>(P value; 95% CI) |
| Female gender | **2.72 (1.13-6.54; P = 0.025)** | **4.72 (1.85-12.02; P = 0.001)** |
| Family history of ARVC/D | N/A |  |
| Family history of VF/SCD | 0.80 (0.18-3.42; P = 0.764) |  |
| Pre-existing VT/VF | 0.89 (0.37-2.18; P = 0.810) |  |
| Syncope | 0.96 (0.41-2.26; P = 0.931) |  |
| Palpitations | 1.06 (0.46-2.42; P = 0.895) |  |
| QRS | 1.01 (0.99-1.02; P = 0.086) |  |
| QTc duration | **1.01 (1.008-1.02; P < 0.001)** | **1.02 (1.01-1.03; P = 0.004)** |
| PR interval | 1.00 (0.99-1.01; P = 0.811) |  |
| R wave amplitude in V5 | 0.69 (0.32-1.49; P = 0.351) |  |
| S wave amplitude in V1 | 0.15 (0.02-1.19; P=0.073) |  |
| Epsilon waves | **4.29 (1.85-9.95; P = 0.001)** | **2.86 (1.21- 6.76; P = 0.017)** |
| PVC | 2.40 (0.88-6.50; P = 0.088) |  |
| TWI in any lead except aVR/V1 | **12.8 (1.72-95.73; P = 0.013)** | **8.36 (1.07-65.3; P = 0.043)** |
| TWI in 2/3 inferior leads | 1.92 (0.84-4.38; P = 0.121) |  |

**Supplementary Table 3.** Cox proportional-hazards model to predict all-cause mortality.

|  | All-cause mortality |  |
| --- | --- | --- |
|  | Univariable Hazards Ratio<br>(P value; 95% CI) | Multivariable Hazards Ratio<br>(P value; 95% CI) |
| Female gender | **3.45 (1.11-10.76; P = 0.030)** | 2.51 (0.54-11.7; P = 0.242) |
| Family history of ARVC/D | N/A |  |
| Family history of VF/SCD | 1.31 (0.29-5.90; P = 0.719) |  |
| Pre-existing VT/VF | 0.63 (0.19-2.06; P = 0.449) |  |
| Syncope | 1.39 (0.49-3.97; P = 0.531) |  |
| Palpitations | 1.73 (0.50-5.28; P = 0.338) |  |
| LVEF | **0.93 (0.89-0.97; P = 0.001)** | **0.89 (0.81-0.97; P = 0.008)** |
| QRS | **1.03 (1.01-1.04; P = 0.001)** | **1.04 (1.01-1.78; P = 0.01)** |
| QTc duration | **1.01 (1.001-1.03; P = 0.029)** | 0.99 (0.98-1.02; P = 0.639) |
| PR interval | 0.99 (0.98-1.00; P = 0.055) |  |
| R wave amplitude in V5 | 0.92 (0.42-2.02; P = 0.834) |  |
| S wave amplitude in V1 | 0.09 (0.006-1.46; P = 0.091) |  |
| Epsilon waves | **4.21 (1.45-12.19; P = 0.008)** | **6.33 (1.16-34.31; P = 0.033)** |
| PVC | 1.93 (0.59-6.37; P = 0.279) |  |
| TWI in any lead except aVR/V1 | 1.53 (0.48-4.91; P = 0.472) |  |
| TWI in 2/3 inferior leads | 1.75 (0.60-5.00; P = 0.304) |  |

**Supplementary Table 4a.** Scoring system for VT/VF Variable Univariable Hazards.

| Variable | Univariable Hazards Ratio (95% CI) | P-value | Points |
| --- | --- | --- | --- |
| Epsilon wave | 2.34 (1.22-4.48; P = 0.011) |  | 1 |
| Syncope | 1.83 (1.04-3.23; P = 0.036) |  | 1 |
| QRS > 98.5 | 2.87 (1.53-5.43; P = 0.001) |  | 1 |

**Supplementary Table 4b.** VT/VF score characteristics of patients with/without incident VT/VF.

|  | With VT/VF | Without VT/VF | P value |
| --- | --- | --- | --- |
| Median risk score for VT/VF (IQR) | 1.0(1.0-2.0) | 0.0(0.0-1.0) | <0.0001 |

**Supplementary Table 4c.** Stratification performance of VT/VF score.

|  | HR (95% CI) | Z value | P-value |
| --- | --- | --- | --- |
| VT/VF score (per unit) | 1.74 (1.30-2.33) | 3.76 | <0.0001 |
| VT/VF score ≥ 1 | 2.08 (1.17-3.7) | 2.49 | 0.013 |

**Supplementary Table 5a.**
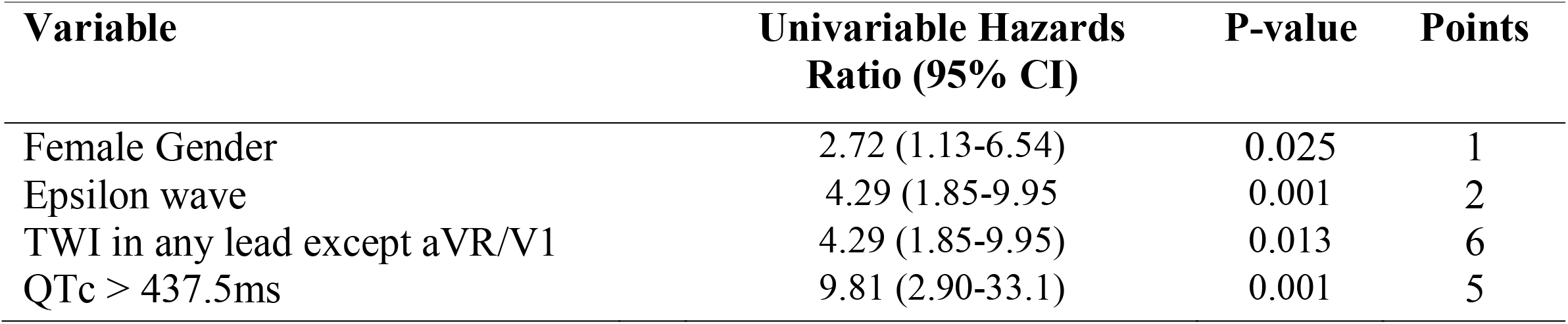
Scoring system for new-onset HFrEF.

| **Variable** | **Univariable Hazards Ratio (95% CI)** | **P-value** | **Points** |
| --- | --- | --- | --- |
| Female Gender | 2.72 (1.13-6.54) | 0.025 | 1 |
| Epsilon wave | 4.29 (1.85-9.95) | 0.001 | 2 |
| TWI in any lead except aVR/V1 | 4.29 (1.85-9.95) | 0.013 | 6 |
| QTc > 437.5ms | 9.81 (2.90-33.1) | 0.001 | 5 |

**Supplementary Table 5b.** HFrEF score characteristics of patients with/without HFrEF.

|  | **Patients with HFrEF** | **Patients without HFrEF** | **P value** |
| --- | --- | --- | --- |
| Median risk score for HFrEF (IQR) | 12 (13-11) | 6 (1-7) | <0.0001 |

**Supplementary Table 5c.** Stratification performance of HFrEF score.

|  | **HR (95% CI)** | **Z value** | **P-value** |
| --- | --- | --- | --- |
| HFrEF score (per unit) | 1.48 (1.26-1.74) | 4.70 | <0.0001 |
| HFrEF score $\geq 11$ | 15.91(5.37-47.17) | 4.99 | <0.0001 |

\*determined by maximal rank statistics approach (Figure 2, left panel)

**Supplementary Table 6a.** Scoring system for all-cause mortality.

| **Variable** | **Univariable Hazards Ratio (95% CI)** | **P-value** | **Points** |
| --- | --- | --- | --- |
| Female Gender | 3.54 (1.11-10.76) | 0.030 | 2 |
| Epsilon wave | 4.21 (1.45-12.19) | 0.008 | 2 |
| QTS > 122.5 | 6.77 (2.34-19.57) | < 0.001 | 3 |
| QTc > 448.5 | 6.05 (1.68-21.8) | 0.006 | 3 |

**Supplementary Table 6b.** Mortalityscore characteristics of patients with/without all-cause mortality.

|  | **Mortality** | **Alive** | **P value** |
| --- | --- | --- | --- |
| Median risk score for mortality (IQR) | 6.5 (5-8) | 2 (0-4) | <0.0001 |

**Supplementary Table 6c.**
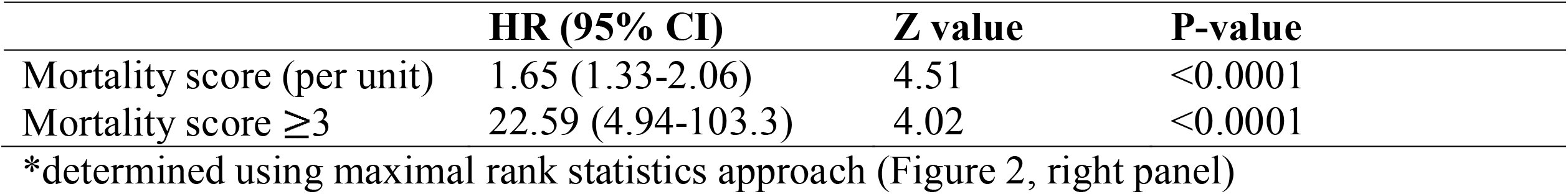
Stratification performance of mortality score.

